# Timing-dependent synergies between noninvasive motor cortex and spinal cord stimulation in chronic cervical spinal cord injury

**DOI:** 10.1101/2025.04.17.25326011

**Authors:** Lynda M. Murray, James R. McIntosh, Jacob A. Goldsmith, Yu-Kuang Wu, Mingxiao Liu, Sean P. Sanford, Evan F. Joiner, Christopher Mandigo, Michael S. Virk, Vishweshwar Tyagi, Jason B. Carmel, Noam Y. Harel

## Abstract

Precise movement requires integrating descending motor control with sensory feedback. Sensory networks interact strongly with descending motor circuits within the spinal cord. We targeted this interaction by pairing stimulation of the motor cortex with coordinated stimulation of the cervical spinal cord. We used separate non-invasive and epidural experiments to test the hypothesis that the strongest muscle response would occur when paired brain and spinal cord stimuli simultaneously converge within the spinal cord. For non-invasive experiments, we measured arm and hand muscle motor evoked potentials (MEPs) in response to transcranial magnetic stimulation (TMS) and transcutaneous spinal cord stimulation (TSCS) in 16 individuals with chronic spinal cord injury (SCI) and 15 uninjured individuals. We compared this noninvasive approach to intraoperative paired stimulation experiments using dorsal epidural electrodes in 38 individuals undergoing surgery for cervical myelopathy. We observed augmented muscle responses to suprathreshold TMS when subthreshold TSCS stimuli were timed to converge synchronously in the spinal cord. At convergent timing, target muscle MEPs increased by 11.0% overall (13.3% in people with SCI, 6.2% in uninjured individuals) compared to non-convergent time intervals. Facilitation correlated with TSCS intensity, with intensity close to movement threshold being most effective. Facilitation did not correlate with SCI level or severity, indicating spared circuits were sufficient for this effect. Noninvasive pairing produced less facilitation compared to intraoperative (epidural) pairing. Thus, sensorimotor interactions in the cervical spinal spinal cord can be targeted with paired stimulation in health and after SCI.

**Highlights:**

- Electrical stimulation of spinal sensory circuits can augment cortical motor evoked potentials, but only when they are timed to arrive synchronously in the spinal cord.
- Facilitation was at least as large in people with spinal cord injury compared with uninjured controls, suggesting that the necessary circuits for this facilitation were spared by injury.
- The magnitude of facilitation with noninvasive facilitation was smaller than the facilitation effect observed with epidural stimulation during elective spinal surgery, but they both rely on precisely timed interactions.
- Facilitation of hand and arm muscle responses correlated with the transcutaneous spinal stimulation intensity, suggesting that the magnitude of the pairing effect can be improved.
- Pairing was effective both in uninjured participants and those with SCI, independently of injury level, severity, or chronicity.

## 1 Introduction

Paired stimulation of the nervous system has been applied in various contexts within humans and animal models to better understand synaptic mechanisms and as a potential method to treat neurological conditions. In the context of improving movement after neural injury, repetitive paired stimulation can target synapses between sensory and motor circuits within the cortex (Stefan et al., 2000, 2002; Wolters et al., 2003), or synapses between sensory, motor, and interneuronal circuits within the spinal cord. Targeting the spinal cord with paired stimulation may consist of coordinated transcranial and peripheral nerve stimulation (Bunday & Perez, 2012; Cortes et al., 2011; Fitzpatrick et al., 2016; Harel & Carmel, 2016; Leukel et al., 2012; Meunier et al., 2007; Shulga et al., 2016; Taylor & Martin, 2009; Yamashita et al., 2021) or coordinated transcranial and spinal cord stimulation (SCS) (Dixon et al., 2016; Knikou, 2017; McIntosh et al., 2024; Mishra et al., 2017; Pal et al., 2022; Pulverenti et al., 2021; Wecht et al., 2021). In all of these approaches, timing plays a critical role, with optimal responses obtained when paired stimulation converges within several milliseconds at target sites.

Paired associative protocols aim to induce *lasting* plasticity through repetitive stimulation. In an effort to optimize the approach, we aim to better understand the potentially facilitatory *immediate* effects of paired stimulation. We have focused on paired stimulation of sensory and motor circuits converging within the spinal cord, specifically the potential of properly timed dorsal stimulation of the cervical cord to amplify upper extremity muscle responses to cortical stimulation (Dixon et al., 2016; Knikou, 2017; McIntosh et al., 2024; Mishra et al., 2017; Pal et al., 2022; Pulverenti et al., 2021; Wecht et al., 2021). This approach may be valuable in amplifying residual descending connections after spinal cord injury (SCI), as even a number of clinically complete SCIs are actually anatomically incomplete (Bunge et al., 1993; A. Kakulas, 1988; B. A. Kakulas, 1999; Sangari et al., 2019). In rodents, subthreshold pulses of epidural stimulation over the dorsal cervical cord enhance the immediate response to electrical motor cortex stimulation by 130-200% when spinal pulses are timed to converge with the arrival of descending cortical pulses (Mishra et al., 2017; Pal et al., 2022). These ‘immediate’ facilitatory effects depend on both cortical descending motor and sensory afferent nerve root transmission. Selective inactivation of either of these pathways fully abrogated the paired stimulation effect in preclinical studies (Pal et al., 2022). In humans undergoing elective cervical spine decompression surgery, paired cortical (using transcranial electrical stimulation, TES) with subthreshold dorsal epidural SCS more robustly enhances immediate arm and hand muscle responses, with effects at least twice as large as observed in rat when timed to converge in the cervical spinal cord (McIntosh et al., 2024). Likewise, suprathreshold dorsal epidural SCS robustly enhances the immediate response to convergent subthreshold TES (McIntosh et al., 2024). Notably, in both rodents and humans, optimal facilitation of motor responses occurs when epidural stimulation occurs over the dorsal root entry zone, slightly lateral to midline, rather than the ventral surface (Greiner et al., 2021; Guiho et al., 2021; McIntosh et al., 2023, 2024; Mishra et al., 2017).

Similar experiments can be performed on awake humans using noninvasive methods. In a demonstration of concept in humans, we showed that subthreshold cervical transcutaneous spinal cord stimulation (TSCS) of the skin overlying dorsal vertebrae can enhance responses to transcranial magnetic stimulation (TMS) of the motor cortex in hand muscles in people with chronic SCI and uninjured participants (Wecht et al., 2021). We designed this study to test the hypothesis in a larger group of people across a wider range of muscles that the strongest effect of paired stimulation would occur when the two stimuli converge within the cervical spinal cord. We compared noninvasive pairing effects to our group’s prior work performing invasive pairing experiments in anesthetized humans undergoing elective cervical spine decompression surgery. This design allows, for the first time, a comparison of non-invasive and invasive methods to test the same hypothesis.

## 2 Material and methods

### 2.1 Participants

All procedures were reviewed and approved by the Institutional Review Board (IRB) of James J. Peters Veterans Affairs Medical Center (JJP VAMC); Weill Cornell Medicine (WCM-IRB, 1806019336); and Columbia University Irving Medical Center (IRB 2, protocol AAAT6563). The study was pre-registered at clinicaltrials.gov (NCT05163639). Written informed consent was obtained prior to study enrollment, and experimental procedures were conducted in compliance with institutional and governmental regulations guiding ethical principles for the participation of human volunteers.

Individuals between the ages of 18 and 80 years living with chronic (> 1 year) cervical SCI and individuals without neurological injury (uninjured individuals) were eligible for recruitment. Individuals with SCI required partially retained motor hand function, scoring 1-4 (out of 5) on manual muscle testing (MMT) of finger extension, finger flexion, or finger abduction in at least one hand. All participants required detectable TMS-evoked motor evoked potentials (MEPs; greater than 50 μV) of the left or right abductor pollicis brevis (APB) or first dorsal interosseous (FDI) muscle. APB was given first preference as the target muscle in each participant. If insufficiently consistent responses were detected in either APB muscle, FDI was given the next preference before the wrist muscles (extensor carpi radialis longus, ECR; flexor carpi radialis, FCR) as alternative target muscles. For participants without neurological injury (uninjured; UI), paired stimulation targeted the dominant arm. For those with SCI, stimulation targeted the arm with lower motor thresholds and more consistent electrophysiological responses to stimulation. Main exclusion criteria included ventilator dependence, open tracheostomy site, open lesions in proximity to stimulating electrodes, use of medications that significantly lower seizure threshold, history of seizures, other significant central nervous system condition, significant coronary artery or cardiac conduction disease, active psychosis, recent history (past 6 months) of uncontrolled autonomic dysreflexia, pregnancy, or implanted electrical or ferromagnetic devices (Rossi et al., 2009, 2021).

### 2.2 General Protocol: Non-invasive experiments

This manuscript reports the results of experiments designed to measure the effects of single or paired stimuli on the immediately ensuing MEP. Experiments were designed to avoid inducing lasting modulation or cumulative effects within the central nervous system. As reported previously (Wecht et al., 2021), participants were seated comfortably, elbows flexed at 90°, hands prone on a pillow, knees extended 80-90°, with feet placed in slight dorsiflexion (0-10°) on a footrest. Wheelchair users had the option of remaining in their chair. A blood pressure cuff (Root® Platform, Masimo Corporation, California, USA) was positioned on the participant’s non-target side. Blood pressure, heart rate, pulse oximetry, and symptoms were recorded frequently during and after each session. Analog-to-digital data acquisition and output systems (National Instruments (NI) USB-6363 and NI USB-6229; Emerson Electric Co., Missouri, USA) controlled with customized LabVIEW software (Emerson Electric Co., Missouri, USA) were used to integrate electromyographic recordings and synchronize stimulator triggers for all experiments.

### 2.3 Electromyographic (EMG) recordings

Skin was prepped (shaved, gently abraded, cleaned with isopropyl alcohol), and surface EMG preamplifiers were placed bilaterally over the APB, FDI, abductor digiti minimi (ADM), FCR, ECR, biceps brachii (BB) short head, triceps brachii (TB) long head, and tibialis anterior (TA) muscles in a belly-tendon montage, held in place with Tegaderm transparent film. Each single bipolar differential preamplifier had built-in x20 gain with internal grounding (MA-411-003 for proximal arm muscles, MA-422-002 for distal hand muscles; Motion Lab Systems (MLS) Inc., Louisiana, USA). A disc ground electrode (Natus Medical Inc., New Jersey, USA) was attached to the lateral epicondyle of the humerus on the target arm. Surface EMG signals were bandpass filtered between 10 and 1000 Hz, sampled at 5000 Hz via an MA400 EMG system (MLS) and two digital acquisition boards (NI USB-6363 and NI USB-6229), and stored for offline analysis.

### 2.4 Transcranial magnetic stimulation

Participants donned an adhesive headpiece with passive markers on the forehead, detected by a stereotactic neuronavigation system (BrainSight; Rogue Research, Montreal, Canada). A MagPro X100 system (MagVenture Inc., Georgia, USA) operating in biphasic mode (anodic-first; 1.0 ms) in conjunction with MagVenture 70 mm butterfly coil (MC-B70) or 80 mm winged coil (D-B80) placed over the motor cortex (M1) hotspot for optimal response in the target muscle. The coil, also fitted with passive markers, was oriented at a 45-degree angle from the medial-sagittal plane so that a posterior-anterior directed electric field perpendicular to the central sulcus was induced in the underlying cortical tissue (Schecklmann et al., 2020).

### 2.5 Transcutaneous Cervical Spinal Cord Stimulation

The top of a single self-adhesive electrode (cathode; 5 cm x 10 cm; UltraStim® X USX2040 rectangular electrode; Axelgaard Manufacturing Co., Ltd., California, USA) was centered vertically over the midline at the identified level of the spinal column that resulted in an MEP at the lowest threshold from the target muscle. This level ranged from C6 to T4. Another self-adhering electrode (anode; same type as the cathode), was placed horizontally over the throat, below the subhyoid depression, roughly two fingers up from the suprasternal notch along the cervicomental angle (CMA)

(Wu et al., 2020). To avoid tracheostomy scar tenderness for one subject and comfort of stimulation in three others, the single anode was replaced by two connected electrodes placed bilaterally over the lateral two-thirds of each clavicle (Einhorn et al., 2013; Murray & Knikou, 2017). Electrodes were held in place with Tegaderm transparent film and connected to a constant current stimulator (DS8R, Digitimer Ltd, Letchworth Garden City, UK). Electrical stimuli were delivered via rectangular constant-current, charge-balanced, biphasic (anodic-first; 1.0 ms/phase) pulses without inter-phase delay.

### 2.6 Motor threshold determination via recruitment curves

During most sessions, resting motor threshold (RMT) at the target muscle hotspot was determined as the stimulation intensity required to evoke a 50 microvolts (μV) motor response in 5 out of 10 repetitions (Rossini et al., 1994). For the final two participants (UI17, SCI17), we transitioned to using a hierarchical Bayesian method of threshold determination (hbMEP) from input-output recruitment curves conducted across a wider range of intensities and muscles (Tyagi et al., 2024). In uninjured individuals, both methods produce comparable RMTs in the target muscle in most cases (average TMS threshold 37.7% of maximal stimulator output (MSO) via 5/10 method, 35.3% MSO via Bayesian method; average TSCS threshold 24.6 milliamperes (mA) via 5/10 method, 24.4 mA via Bayesian method). In addition to the 5/10 method, we defined the highest stimulus intensity without visible real-time responses (zero responses out of 10 repetitions) via an oscilloscope (DPO2014B, Tektronix Technology Co. Ltd., Shanghai, China). We referred to this stimulus intensity as ‘TMS_000’ or ‘TSCS_000’ for TMS and TSCS, respectively.

Input-output recruitment curves in response to brain stimulation were assembled via delivery of single pulses at 0.125 (8 seconds) or 0.2 Hz (5 seconds) at varying intensities in pseudorandom order ranging from subthreshold intensities to 97% MSO, or the highest tolerable intensity. Similarly, recruitment curves in response to single-pulse spinal stimulation (0.125 - 0.2 Hz) ranged from subthreshold intensities to 136 mA, or the highest tolerable intensity. Recruitment curves for participants SCI 1 through 7 and UI 1 through 11 were assembled with 8 repetitions at 6 to 9 discrete intensities relative to 5/10 method RMT. Recruitment curves for the remaining participants were assembled with one repetition at 48 discrete intensities ranging between 0 and maximal tolerated intensity. All recruitment curves were analyzed post hoc using the hierarchical Bayesian approach to calculate the Bayesian method RMT (Tyagi et al., 2024). Note that in some cases, this resulted in reported spinal stimulation intensities above the Bayesian threshold (Table 1), even though at the time of experimentation, they were below threshold using the 5/10 method.

**Table 1.**
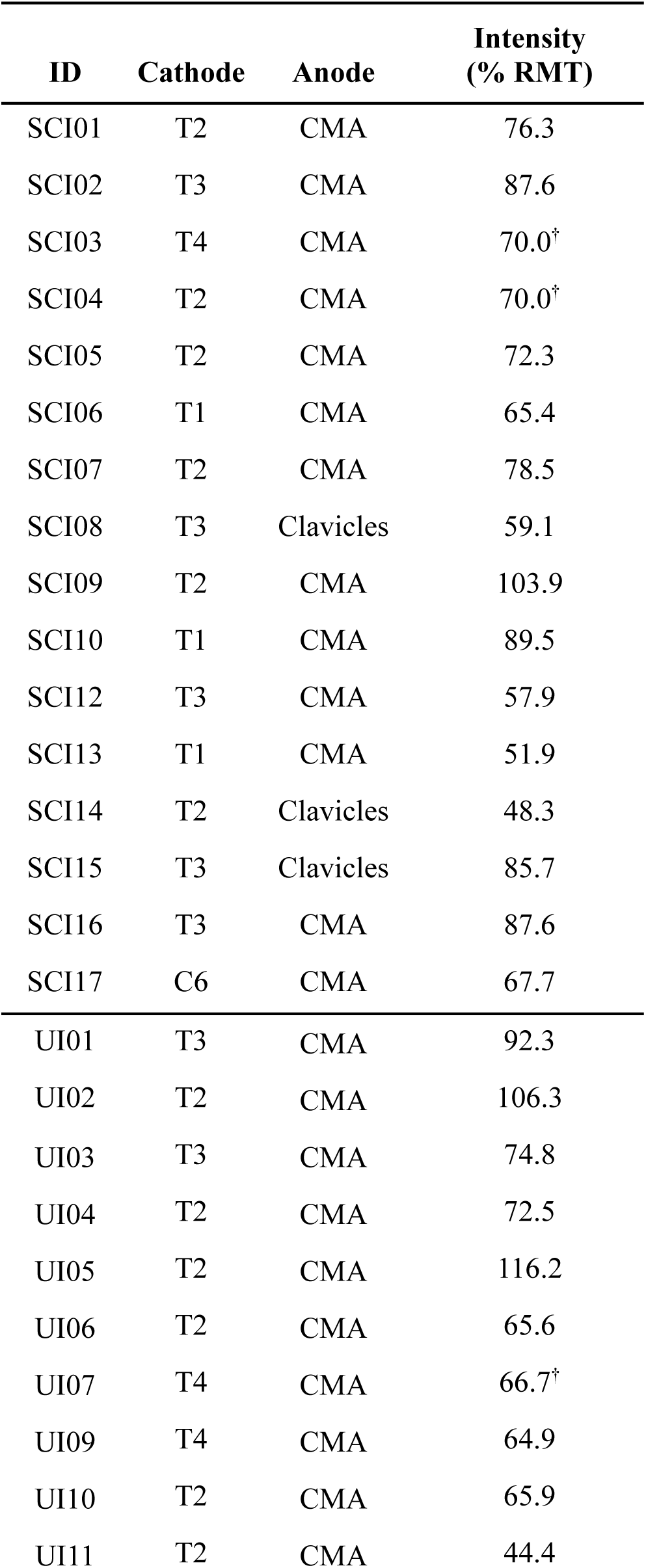
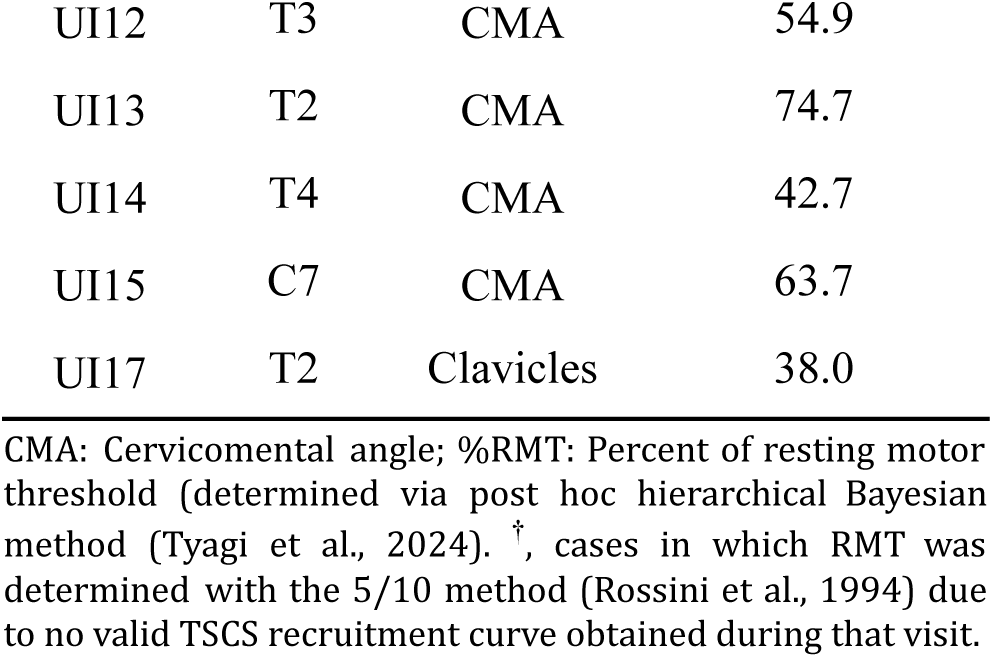
Spinal stimulation characteristics.

### 2.7 Immediate Effects

All participants underwent immediate effects experiments to test the interaction between TMS and TSCS across a range of pairing intervals (PIs). To compare results among participants with variations in central and peripheral nerve conduction velocities, individualized PIs were determined according to calculated stimulus arrival in the cervical spinal cord. A PI of ‘0’ represents the simultaneous arrival of TMS and TSCS within the spinal cord. Positive PI values represent TMS arrival before TSCS. Negative PI values represent TSCS arrival before TMS (Fig. 1B). Given that the site of TSCS excitation occurs at spinal nerve roots at a latency of roughly 1.6 ms from motor neuron cell bodies (Mills and Murray 1986), peripheral motor conduction time (PMCT) was calculated by adding 1.5 ms to the target muscle response latency when TSCS pulses were delivered at 200% of RMT (Dixon et al., 2016; Pulverenti et al., 2019). Central motor conduction time (CMCT) was calculated by subtracting PMCT from the target muscle response latency when TMS pulses were delivered at 120% of RMT (Wecht et al., 2021). Thus, to achieve a PI of 0, a spinal stimulus would need to be delivered with a delay of (CMCT - 1.5 ms). Prior to the final analysis of the accumulated dataset, all individual pulse response latencies were manually reviewed for accuracy while blinded to pairing conditions. This led to several adjustments to individual CMCTs and corresponding PIs averaging 0.27 ms.

**Figure 1.**
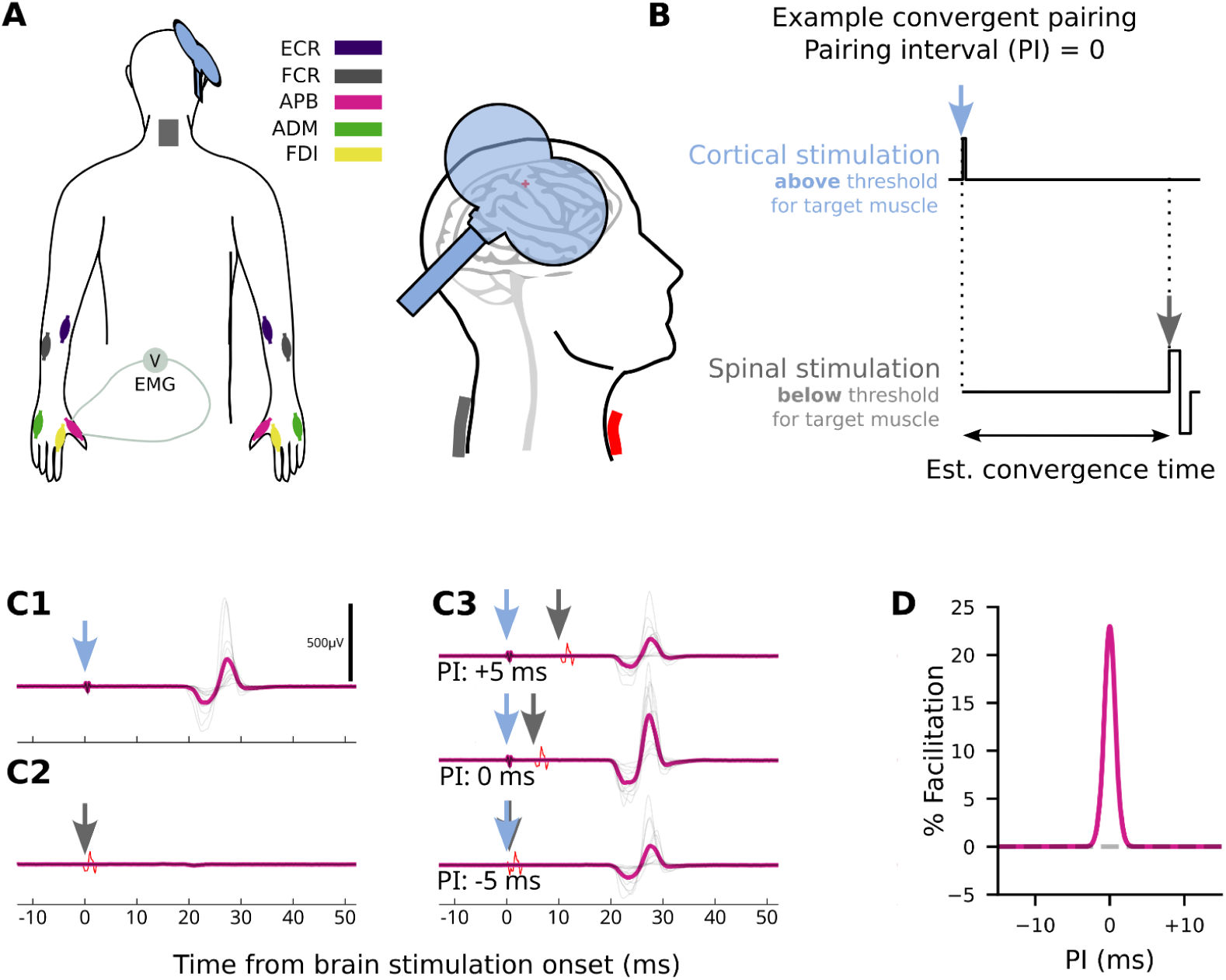
Transcutaneous spinal cord stimulation and transcranial magnetic brain stimulation experiment. **A,** Schematic of electromyography (EMG) and stimulator placement. Colors correspond to different analyzed muscles. Transcranial magnetic stimulation (TMS) was positioned over the target muscle ‘hotspot’. Transcutaneous spinal cord stimulation (TSCS) cathode electrode shown centered over the T1 spinal level. ADM, abductor digiti minimi; APB, abductor pollicis brevis; ECR, extensor carpi radialis; FCR, flexor carpi radialis; FDI, first dorsal interosseous. **B,** The timing between pulses of suprathreshold TMS delivered over the motor cortex, and pulses of subthreshold TSCS delivered over the cervical spine is varied such that pulses converge at the cervical spinal cord at different intervals relative to the estimated central motor conduction time. Example shown where pulses converge simultaneously (Pairing Interval (PI) = 0). Blue and gray arrows indicate cortical and spinal stimulation, respectively. **C,** a subset of example MEPs including brain-only and spinal-only conditions. Blue and gray arrows as in B. **C1,** Response to brain-only stimulation at 120% of the experimentally determined resting motor threshold for the target muscle. **C2,** Response to subthreshold spinal-only stimulation. **C3,** Responses to paired stimulation. Averaged responses at different PIs show that the 0 ms PI response appears larger than responses at non-convergent times. **D,** Quantification of pairing facilitation in a single participant. A bell-shaped curve is fitted to MEP size data. The magnitude of the peak or trough relative to the offset of the fitted curve quantifies the estimated facilitation or depression, respectively

During pairing experiments, pulses of TMS were delivered either at suprathreshold (120% of TMS_RMT) or subthreshold (90% of TMS_RMT or TMS_000) intensity. Pulses of TSCS were delivered at subthreshold intensity. Note that for the first participant with SCI and the first two uninjured individuals, a set percentage (90%) of the threshold determined via the 5/10 method was used. Due to an excessive number of individual suprathreshold responses to stimulation at this intensity, we further reduced TSCS intensity to 70% of the 5/10 method threshold in six SCI and four uninjured participants. Thereafter, we chose a spinal stimulation intensity level that would result in zero responses out of 10 repetitions. See Table 1 for stimulus intensities used as a percentage of Bayesian RMT. Pairing intervals ranged from -50 ms to + 30 ms. In one experiment, paired or unpaired stimuli were delivered at 0.1 Hz (10 seconds) in pseudorandomized order for a total of 12 repetitions per condition (22 paired, 3 unpaired conditions) for a total of 300 stimulation events. For participant comfort, the 300 events were divided into three runs of 100 events (each taking roughly 17 minutes). Each run was separated by several minutes. Four SCI participants (SCI10, SCI12, SCI13, SCI16) underwent a narrower range of PIs focused on combinations of suprathreshold TMS with subthreshold TSCS, delivered at 0.1 Hz in pseudorandomized order for a total of 12 repetitions per condition (9 paired, 2 unpaired conditions) for a total of 132 stimulation events. These were divided into two runs of 66 events (each taking roughly 11 minutes).

### 2.8 Data Analysis

#### 2.8.1 MEP size calculation

MEPs were quantified using the area under the curve (AUC). In order to suppress SCS artifact, raw MEPs from each muscle were linearly interpolated within a 5 ms window time-locked to the spinal stimulation (see example in Fig. 1C3, red). In all pairing interval experiments, EMG AUC was measured in a window between 5 ms and 90 ms after the TMS trigger, except for unpaired TSCS, in which AUC was measured in the same window after the TSCS trigger.

#### 2.8.2 Response, run, and muscle rejection

We built an analytical model to quantify the effects of pairing subthreshold SCS with suprathreshold TMS based on their relative convergence in the spinal cord. We focused on the distal musculature in the arm targeted by TMS because we found the muscles of the upper arm to be more susceptible to spinal stimulation artifact. We analyzed a total of 11,746 stimulation events (378±73 per participant) for a total of 55,109 MEPs. Of these stimulation events, 8,306 (268±61) were performed during brain-spinal cord pairing experiments and used to assess the pairing effect. 3,440 (111±33) were performed during single-modality recruitment curve collection and used for threshold estimates. Due to slight variations in recording layout across participants, on average we recorded 4.7 of 5 distal MEPs per stimulation event. FDI was not recorded in 2 SCI and 5 uninjured participants.

We analyzed all paired stimulation data with valid trigger timings above PIs of -15 ms to ensure that MEPs generated by TSCS were consistently present in the MEP size calculating window. Individual runs/muscles were excluded based on the following criteria: 1) stimulation artifact that persisted beyond the interpolation window, 2) response that was totally absent for a run, and 3) systematic discontinuity in the MEP size over time. With these criteria, 99.98% of target muscle run data and 91.8% of all muscle MEPs were analyzed.

#### 2.8.3 MEP size models

##### 2.8.3.1 Statistical model of paired stimulation for the target muscle

To quantify the effects of paired stimulation, we fitted a curve to model facilitation as a function of PI. We observed a bell shape to this curve in our previous experiments using epidural paired stimulation in rats (Mishra et al., 2017) and in the intraoperative experiments in people (McIntosh et al., 2023). Fitting a curve allows us to combine data with different PIs across participants.

First, MEP size was log-transformed to approximate normality(Nielsen, 1996). For an individual run, the transformed MEP size observation was modeled as a normally distributed random variable 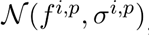 where indices represent intensity and participant, respectively. The expected value (*f* consists of a stimulus-dependent modulation-based function (*g* of the pairing interval as well as baseline components that tracked the MEP size in the absence of facilitation: 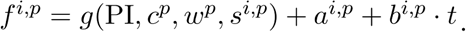 The observation noise was distributed as 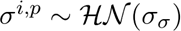 where 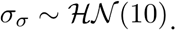

The bell-shaped function (g) was defined as: 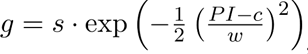 dependent on the peak size (*s*), peak center (*c*, an adjustment of the PI), and peak width (*w*). The offset (*a*) and coefficient of the linear term in time (*b*) account for the expected local baseline MEP size during an experiment in the absence of facilitation (i.e. when *g*(.) = 0.

Statistical modeling was performed with hierarchical Bayesian methods. The primary outcome measure was facilitation of TMS MEPs with paired suprathreshold TMS and subthreshold TSCS. Priors were specified to account for variability across intensities and participants. Individual participant estimates of the primary outcome measure were modeled as 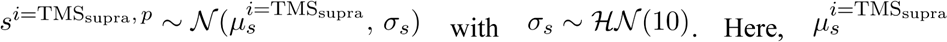 is the population-level facilitation parameter, representing the central tendency from which individual participant facilitation estimates are drawn. A positive shift of this parameter away from its weakly informative unbiased prior distribution *N*(0,10)) indicates facilitation at the population level, while a negative shift would indicate suppression. Hypothesis testing was conducted by assessing whether the probability of 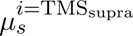 being greater than zero is over 95%. The same model structure applied for *i* = TMS_sub_, the subthreshold TMS condition.

Parameters *c*^*p*^ and *w*^*p*^, which govern facilitation characteristics beyond peak size, were participant-specific. The peak center parameter *c*^*p*^ was assigned a hierarchical prior centered at zero: 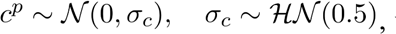 with *c*^*p*^ bounded between -2 and 2. Informative priors for *c* were chosen because: 1) the optimal pairing interval (PI = 0) was experimentally determined based on the individual participant MEP onset timings, and 2) they improved model convergence, likely because in cases of minimal individual facilitation, any value of *c* is a possible solution. The width parameter *w*^*p*^ was bounded to positive values between 0.5 and 5.0, drawn from a truncated normal distribution 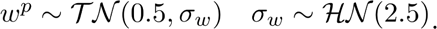 The rationale for the assumptions on *w*^*p*^ are based on our previous observations from intraoperative experiments in people (McIntosh et al., 2023) and to improve model convergence as for *c*^*p*^.

To extend the model to capture multiple runs within an experiment, we extended the offset parameter to be indexed by the run and modeled their relationship to each other as Laplace distributions, 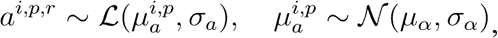 reflecting variability across participants and visits but stability within experiments. The following hyperparameters were used: 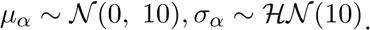 Observed drift in MEP size over the duration of each run was modeled using a coefficient of linear term in time *b*^*i,p,r*^, with the time variable reset to zero at the start of each run. The hierarchical structure for this parameter was the same as for the offset term (*a*).

##### 2.8.3.2 Calculation of % facilitation

MEP size facilitation at the optimal pairing *M*^*o*^ relative to the MEP size at the sub-optimal pairing *M*^*n*^ was expressed as a % change as follows: 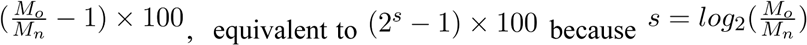 due to the MEP size modeling taking place on a log base 2 scale. Similarly, the population-level facilitation was calculated as (2^μs^ − 1) × 100.

##### 2.8.3.3 Extension of the statistical model of paired stimulation for multiple muscles

In order to extend the model to capture all muscles rather than only the target muscle, we introduced separate parameters for each muscle. At the lowest level of the hierarchy, facilitation parameters (*s, c, w*) as well as baseline parameters (*a, b*) were drawn from muscle-level hierarchical priors. An observation mask was used to seamlessly deal with missing data in specific muscles.

##### 2.8.3.4 Extension of the statistical model of paired stimulation for cohort descriptor (UI versus SCI)

To address whether both the UI and SCI groups exhibited facilitation and investigate potential differences between the two groups, facilitation was extended to be captured by 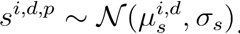 Here, μ^i,d^_s_ is the population-level facilitation parameter for the different TMS intensity conditions (*i*) as well as the different cohort descriptor (absence/presence of SCI) (*d*).

##### 2.8.3.5 Recruitment curve fitting

Recruitment curves were estimated using hbMEP (Tyagi et al., 2024) with a rectified logistic curve and a mixture extension of the standard hierarchical Bayesian model. The hyperpriors of the model were adjusted to estimate curves for both TSCS and TMS data using the same model on a similar intensity scale (see Supplementary Fig.1). Missing data in specific muscles was handled with an observation mask. The model yielded posterior distributions of the curve parameters, including threshold, and the mean of these posteriors were used as point estimates for subsequent analyses. To avoid using highly uncertain threshold estimates, we discarded estimates with a 95% highest density interval (HDI) (Kruschke, 2014) greater than 30 for both TMS or TSS recruitment curves. This resulted in the omission of a single TMS threshold estimate where the target muscle was not activated within the tested stimulation intensity range.

##### 2.8.3.6 Recruitment curve fitting with dependence on the presence of SCI

A group comparison model from hbMEP was used to compare the thresholds of groups of SCI and uninjured participants on TSCS and TMS data. For each group of participants, the model yielded a posterior distribution of their thresholds. In this comparison of groups, the 95% HDI of the difference between the threshold posterior of SCI and uninjured participants is reported, and statistical significance was concluded if the HDI excluded zero.

#### 2.8.4 Secondary statistics

Values are reported as mean ± standard error of the mean (SE) or standard deviation (SD) unless otherwise stated. To assess the relationship between pairs of variables, linear model fits were performed using linear regression, and the associated slope, *p*-value, and coefficient of determination *R*^2^ were also reported.

#### 2.8.5 Code version and availability

Pre-processing was performed with MATLAB, R2024a (MathWorks Inc., Natick, MA, USA). Subsequent analysis was performed with Python (v3.11). Bayesian models were estimated with NumPyro’s (v0.13.2) implementation of the No-U-Turn (NUTS), making use of hbMEP (v0.6.2) for recruitment curves. Analysis and model implementation code is publicly available at https://github.com/jrmxn/brain-spinal-pairing.

### 2.9 Intraoperative data and analysis

In order to compare the effect of non-invasive pairing of brain and spinal stimulation on MEP size facilitation to intraoperative facilitation, as well as to validate the developed facilitation model, we re-analyzed previously published data from a single-session physiology study. Detailed protocols have been previously described (McIntosh et al., 2024). Relevant sections are reproduced here. Participants were adult patients with cervical spondylotic myelopathy and/or multilevel foraminal stenosis requiring surgical intervention. Patients were enrolled from the clinical practices of the spine surgeons participating in that study.

For epidural SCS, flexible catheter electrodes placed on the dura were used for stimulation during clinically indicated surgeries. A single biphasic pulse (pulse-width = 250 μs) was used for stimulation. Motor cortex stimulation was performed with TES. A monophasic triplet pulse with a width of 75 μs and a pulse separation of 3 ms was delivered through subdermal needle electrodes placed in a quadripolar montage at C1, C2, C3 and C4. The two subdermal electrodes in the hemisphere targeted for stimulation served as the anode, and the two electrodes on the opposite hemisphere served as the cathode. Surface EMG recordings were taken from muscles selected as per standard of care, with additional recordings from wrist muscles. Recordings were made at a sampling rate of 8.3 kHz and band-pass filtered between 10 Hz and 2 kHz.

Paired stimulation was performed with TES delivered at 110% of MEP threshold and subthreshold spinal stimulation at 90% of MEP threshold for a targeted muscle. The interstimulus interval (ISI; n.b. not the pairing interval) between the initiation of TES and subsequent epidural SCS was varied between 3 and 13 ms.

For the purposes of the subsequent analysis, to match the non-invasive analysis, MEPs were quantified in hand and wrist muscles ipsilateral to the side of stimulation, with the rectified AUC calculated in a window between 6.5 ms and 75 ms after the start of the first stimulation pulse. This analytic model produced results that are more conservative per individual than the prior intraoperative analysis. The same target muscle and multi-muscle facilitation model were used consistent with the non-invasive data, with the following exceptions:

1. During intraoperative experiments, tested pairing intervals were not adjusted dependent on the CMCT estimate from MEP onset times. To analyze the intraoperative data relative to the estimated PI = 0, consistent with the non-invasive data analysis, we consequently subtracted our previous estimate of optimal pairing ISI based on the difference between cortical and spinal MEP onset times (see Figure 4 of (McIntosh et al., 2024)) from the tested ISIs. This results in the maximum facilitation occurring close to a pairing interval of zero. Because the previous estimate could only be made for 14 participants, we chose to average these values and relied on the model to estimate the *c*^*p*^ rather than correcting each participant individually. To account for these differences in the data, the peak center parameter (σ_*c*_) was drawn from *HN*(1.0), where the hyperprior parameter setting was estimated from the differences between calculated optimal pairing ISI from MEP stimulus onset times versus from facilitation peak timing.
2. Experiments typically lasted less than 5 minutes, and we did not observe substantial drift. The coefficient that allows the baseline MEP size to vary over time (*b*) was consequently excluded from the model.

## 3 Results

### 3.1 Participant recruitment and characteristics

Sixteen individuals with chronic SCI and 15 uninjured individuals without central nervous system injury participated in the noninvasive study (Table 2). The SCI group included 13 males and 3 females, whereas the uninjured group included 8 males and 7 females. The average age of those with SCI was 47.4 ± 18.3 years, and of those without injury was 31.5 ± 10.1 years. Participants with SCI had motor levels ranging from C2 to C8, with total upper extremity motor scores averaging 32.4 ± 10.3 (range 14-47 out of 50), specifically 16.8 ± 5.1 (out of 25) for the targeted side. Among those with SCI, the target muscle (the hand or wrist muscle with the most consistent TMS thresholds) was APB in 11, FDI in 4, ECR in 3, and FCR in 1. For all uninjured participants except one (FDI), the target muscle was APB.

**Table 2.**
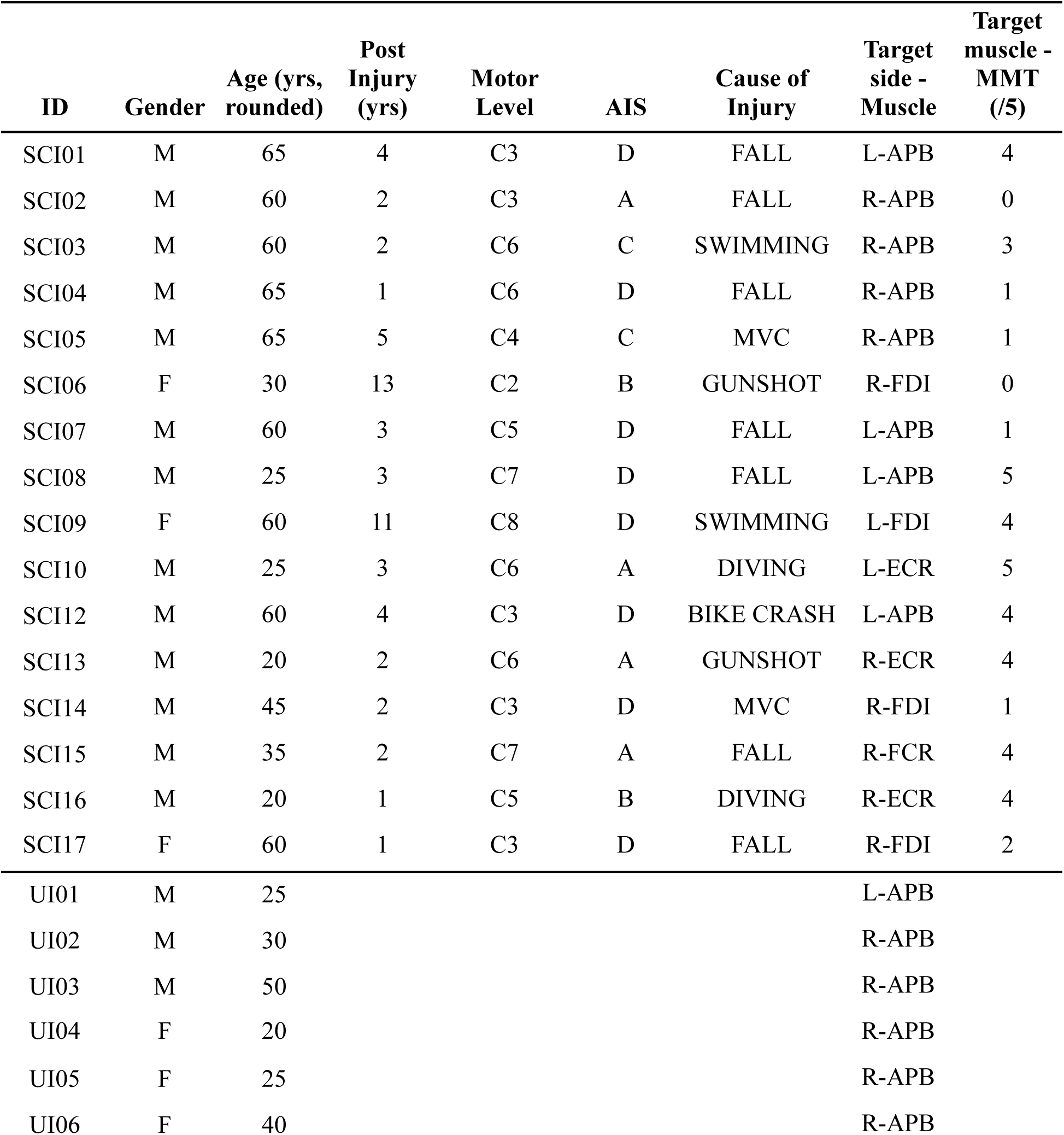
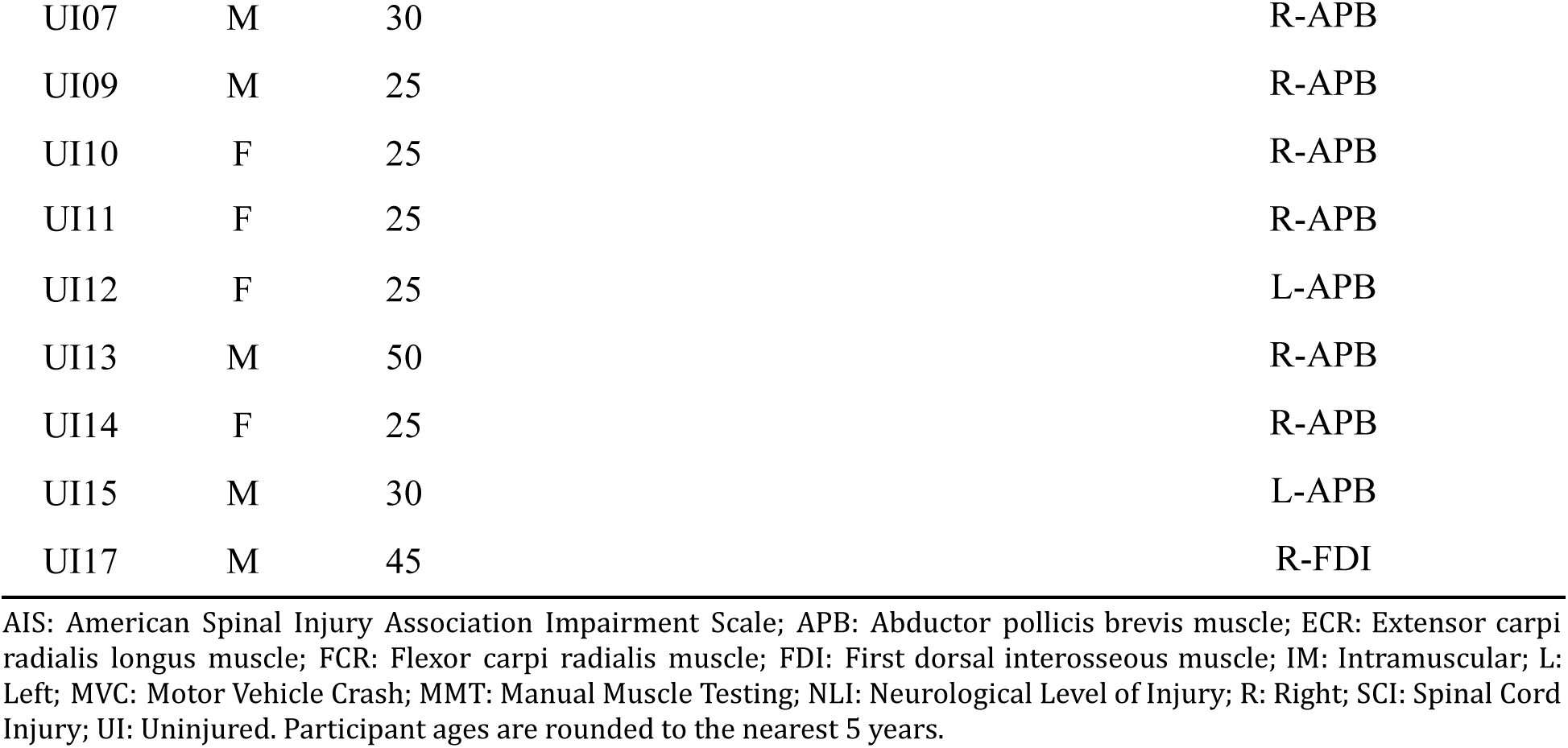
Participant Demographics.

### 3.2 TSCS augments motor cortex MEPs at the predicted convergence time

We used MEP size to quantify the effects of pairing subthreshold TSCS with suprathreshold cortical stimulation. Pairing between TMS and TSCS was calibrated to each participant’s CMCT, as detailed in Methods section 2.7. In brief, a PI of 0 represents the simultaneous arrival of TMS and TSCS within the cervical spinal cord. Positive PI values represent TSCS arrival after TMS. Negative PI values represent TSCS arrival before TMS. PIs between -10 ms and +10 ms were more extensively tested.

As noted in Fig. 2A, convergently timed subthreshold TSCS facilitated target muscle MEP responses to suprathreshold TMS by 11.0% on average when the stimuli arrived at the same time in the spinal cord (PI = 0). The facilitatory effect was robust, with a posterior probability of 99% using a Bayesian model. APB was the most commonly targeted muscle, showing an average facilitation of 9.8% but a high degree of variability (Fig. 2B). FCR and APB muscles had the most consistently positive facilitatory responses at PI = 0. In contrast, pairing subthreshold TMS with subthreshold TSCS did not result in facilitation in the target muscle (0.4% on average, with a posterior probability of 47%). Together, these results indicate a highly significant but moderate pairing effect between suprathreshold TMS and subthreshold TSCS that was observed in hand and wrist muscles across people with and without SCI.

**Figure 2.**
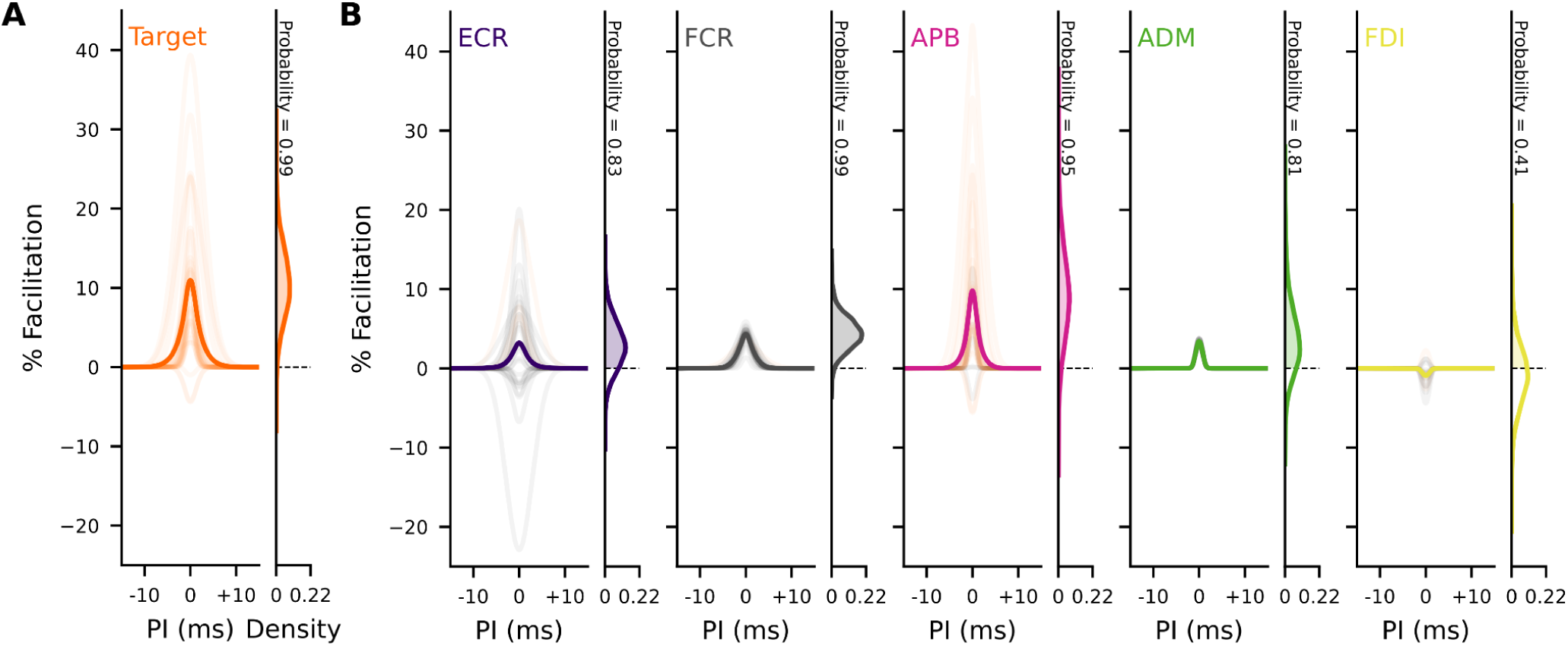
Pairing non-invasive brain and spinal stimulation produces facilitation at the spinal convergence time. The left side of each panel shows the percentage facilitation of motor evoked potential responses to transcranial magnetic stimulation (TMS) when transcutaneous spinal cord stimulation (TSCS) pulses arrive at the spinal cord at pairing intervals (PIs) ranging from 15 ms before to 15 ms after TMS pulse arrival. The right side of each panel shows the probability distribution of the population level facilitation parameter. Probability of % Facilitation > 0 indicated. Bold lines indicate average results for the target muscle. Faint lines indicate the results of individual participants. **A.** Convergent pairing of TMS and TSCS facilitates the targeted muscle across all participants (PI = 0 is used to align maximum facilitation). **B.** Results for each muscle across participants. Facilitation was found to be consistently positive in FCR, APB and ADM but not ECR or FDI. ADM, abductor digiti minimi; APB, abductor pollicis brevis; ECR, extensor carpi radialis; FCR, flexor carpi radialis; FDI, first dorsal interosseous.

### 3.3 Pairing facilitation depends on stimulation intensity and spinal threshold

We hypothesized that spinal stimulation intensity and the baseline responsiveness of target muscles to TMS would predict facilitation. In one SCI and two uninjured participants, we delivered spinal stimulation at a set level of 90% of threshold. In six SCI and four uninjured participants, we delivered spinal stimulation at a set level of 70% of threshold.

In other pairing experiments, we delivered spinal stimulation at intensities that resulted in zero responses out of 10 repetitions of unpaired TSCS (Table 1). All thresholds were recalculated post hoc using the hbMEP technique (Section 2.6), resulting in stimulation intensities used during pairing that ranged from as low as 38% of hbMEP threshold to as high as 116% of threshold, averaging 71.7% ± 3.5% of threshold (Fig. 3A2). We predicted that higher intensities relative to threshold would result in a larger pairing effect. The magnitude of pairing facilitation indeed correlated with increasing spinal stimulation pairing intensity in the target muscle (*p* = 0.03, R^2^ = 0.17, slope = 0.19; Fig. 3A3). Conversely, pairing facilitation did not correlate with cortical TMS intensity or threshold (*p* = 0.21, R^2^ = 0.07, slope = 0.13; Fig. 3B).

**Figure 3.**
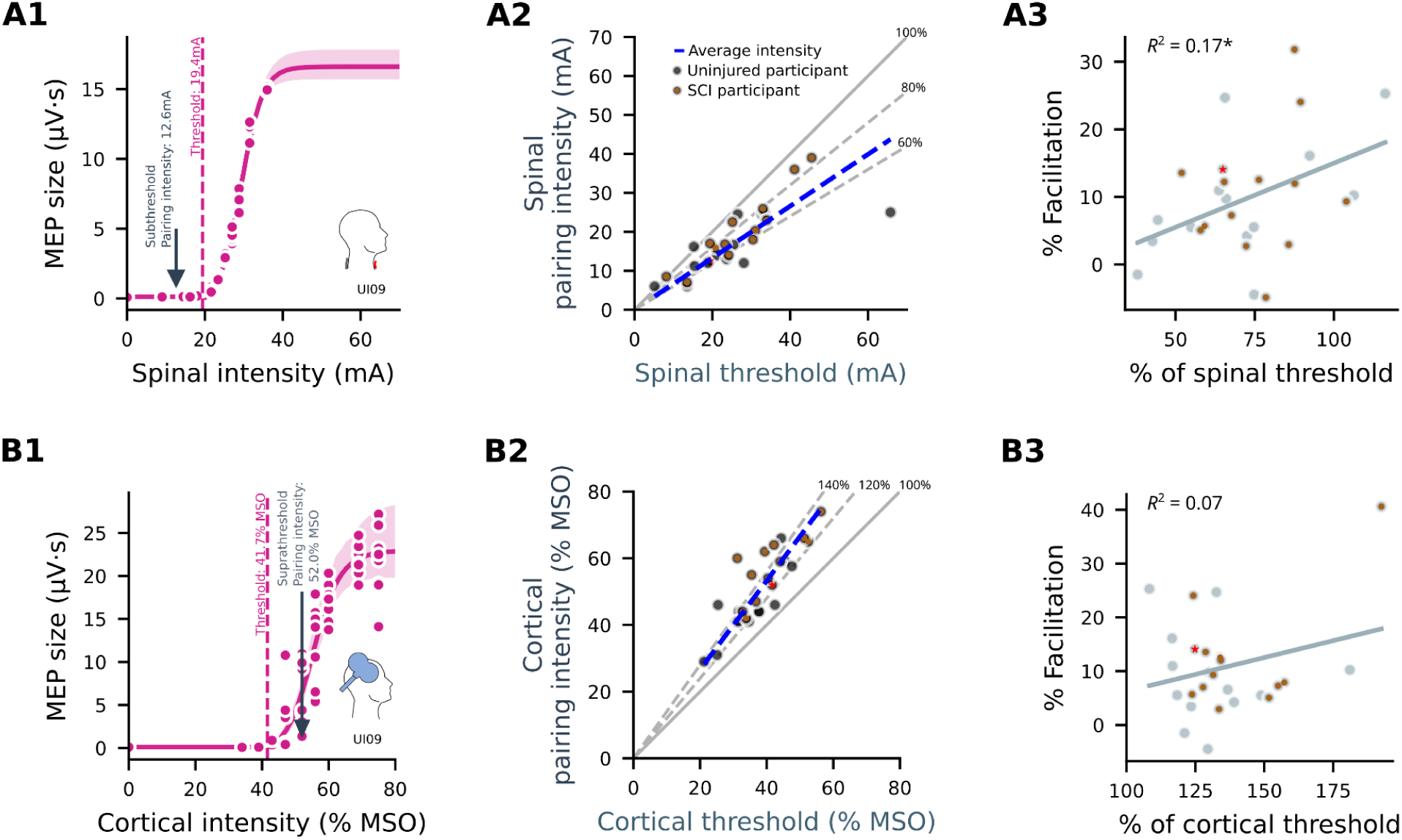
Predictors of pairing facilitation for the targeted muscle. **A.** Pairing facilitation correlates with spinal stimulation intensity. **A1,** Example spinal recruitment curve showing estimated curve fit and mean threshold estimate (dashed line) as well as the spinal stimulation intensity used for pairing (arrow). **A2,** Spinal stimulation intensity used during pairing versus the spinal threshold determined by curve fitting. Dashed blue lines represent linear fit, indicating stimulation intensity as a percentage of the threshold. Brown dots mark participants living with spinal cord injury (SCI). **A3,** Facilitation is greater in individuals receiving higher spinal stimulation intensity as a percentage of threshold. * indicates statistically significant (p < 0.05). **B1-B3,** Analogous data for suprathreshold cortical stimulation. mA, milliamperes; MSO, maximal stimulator output.

**Figure 4.**
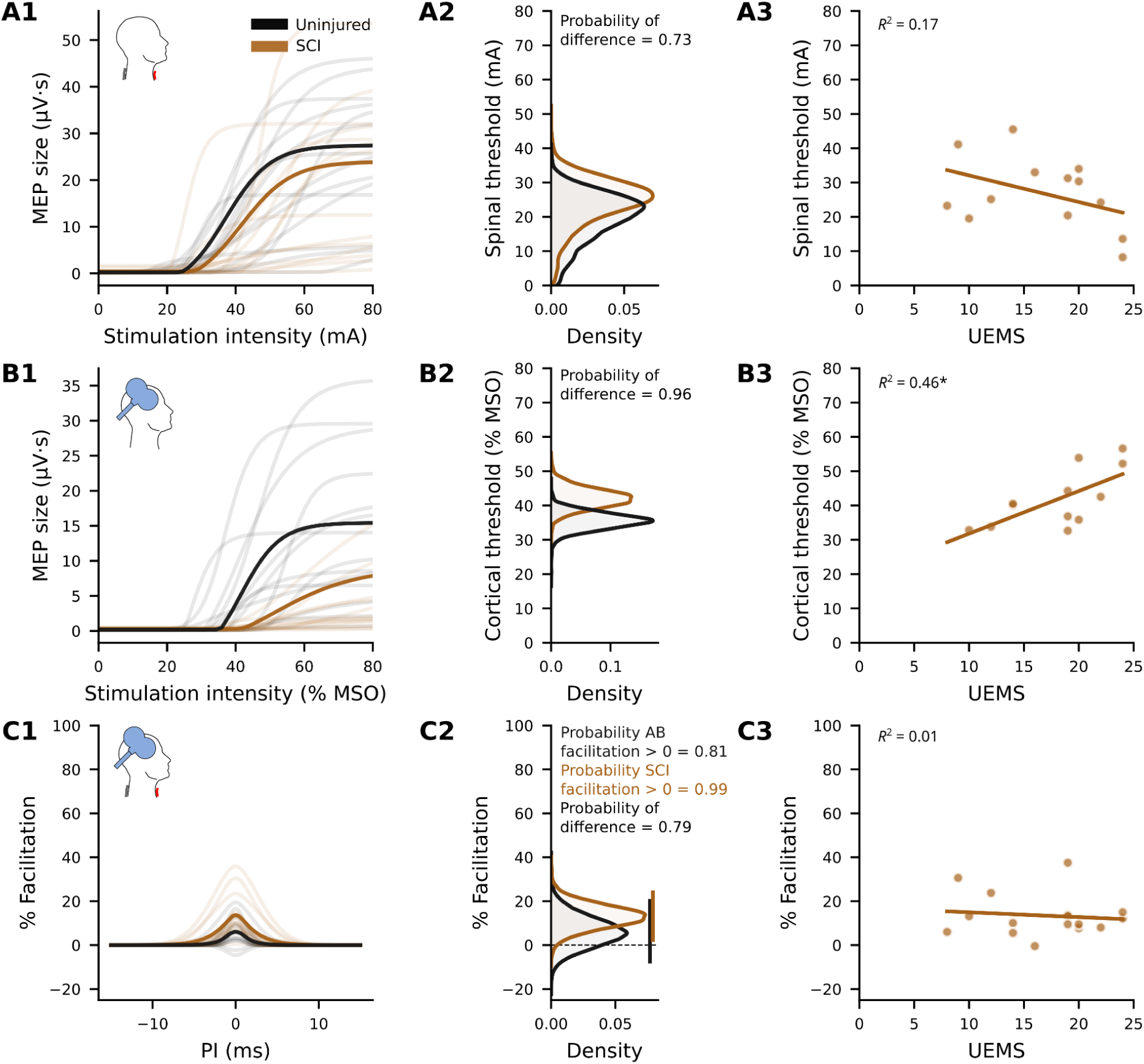
Uninjured participants and participants living with spinal cord injury (SCI) differ in cortical thresholds but not spinal thresholds or pairing facilitation. **A1:** Uninjured and SCI participant spinal stimulation recruitment curves for the targeted muscle. Solid lines represent recruitment curves generated from the averaged parameters of uninjured (black) and SCI (brown) participants. Light-shaded lines represent individual participants. **A2:** Population level threshold distributions for spinal stimulation do not show a clear separation between groups. **A3:** Spinal thresholds show a non-significant inverse correlation with the target arm’s upper extremity motor score (UEMS) in SCI participants. **B1:** as for A for cortical stimulation. **B2:** Thresholds for cortical stimulation separate, indicating higher thresholds for SCI than for uninjured participants. **B3:** Cortical thresholds increase with increasing UEMS. **C1:** Uninjured and SCI facilitation curves for the targeted muscle. Solid lines represent the magnitude of facilitation averaged across uninjured (black) and SCI (brown) participants. Light-shaded lines represent individual participants. **C2:** Population level facilitation is positive for both uninjured and SCI participants, but evidence that facilitation is greater than zero is only strong for SCI participants and not for uninjured participants. **C3:** Facilitation does not correlate with UEMS. mA, milliamperes; MEP, motor evoked potential; MSO, maximal stimulator output; PI, pairing interval.

### 3.4 Uninjured participants and those with SCI differ in cortical thresholds but not spinal thresholds or pairing facilitation

TSCS thresholds in the target muscle did not differ between groups (Fig. 4), as expected, because the cathode was placed caudal to the level of injury (Methods 2.5). TSCS threshold tended to decrease with increasing target arm motor score (*p* = 0.16, R^2^ = 0.17, slope = -0.78). Also, as expected, participants with SCI had higher TMS thresholds than uninjured individuals (Fig. 4). Surprisingly, TMS thresholds increased with increasing target arm motor scores (*p* = 0.02, R^2^ = 0.46, slope = 1.24). In terms of pairing facilitation, participants with SCI facilitated to a greater degree on average (13.3% versus 6.2% in uninjured individuals), but the difference across groups was not significant (Fig. 4C). Only participants with SCI showed an overall facilitation greater than 0. Importantly, facilitation did not correlate with the target arm motor score (*p* = 0.67, R^2^ = 0.01, slope = -0.22; Fig. 4C3) or participant age, injury level, or injury duration (Supplementary Fig.2).

### 3.5 Intraoperative pairing experiments validate the model-based approach and demonstrate much greater facilitation than non-invasive pairing

We compared results obtained using non-invasive pairing against results of paired stimulation conducted during clinically indicated surgeries using epidural SCS. These experiments, which have been described before (McIntosh et al., 2024), were newly analyzed with the same modeling described in Sections 2.8 and 2.9 to be able to compare the results directly with the non-invasive experiments. As shown in Fig. 5, pairing of TES with epidural SCS induced facilitation of 400% compared with 11% facilitation in non-invasive pairing. This facilitation was greatest in hand muscles. Thus, paired stimulation, which requires precise timing to induce facilitation, can be induced non-invasively, though to a lesser degree than epidural stimulation.

**Figure 5.**
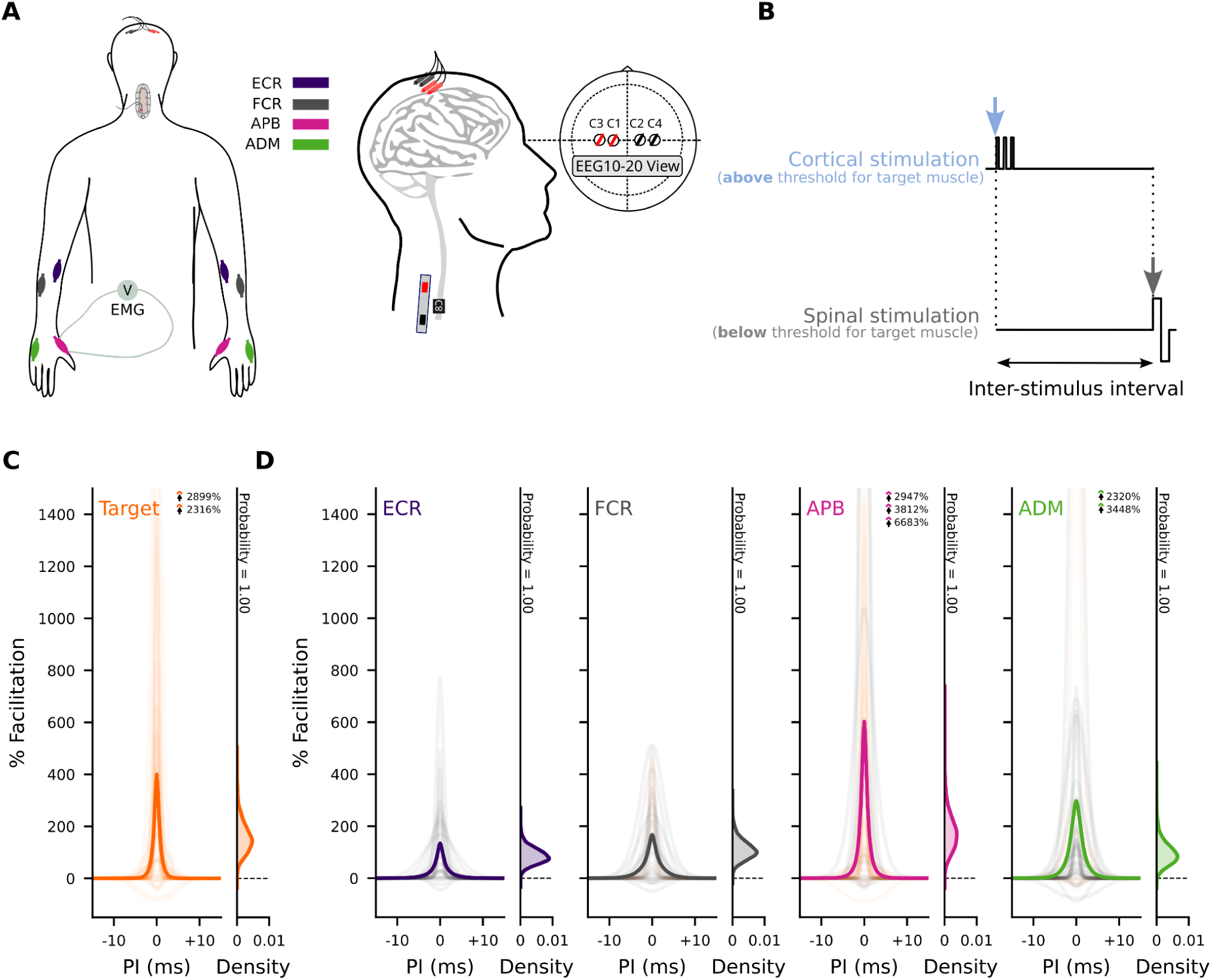
Intraoperative pairing experiments demonstrate strong facilitation. **A**, colors correspond to different analyzed muscles (see legend). Subdermal needles were placed for transcranial electrical stimulation (TES) of the brain using the EEG 10–20 system (red represents anodes, black represents cathodes). Epidural spinal cord stimulation (SCS) electrode shown placed below the lamina on the posterior aspect of the spinal cord. Adapted from (McIntosh et al., 2023). **B**, three pulses are delivered over the motor cortex, followed by a variable inter-stimulus interval (ISI) before a single pulse is delivered to the spinal cord. Tested ISIs were determined to include the estimated convergence time (PI = 0, which is also used to align maximum facilitation). **C**, Facilitation analyzed for intraoperative data using the same model consistent with the non-invasive data. Target muscle shows strong evidence for facilitation, consistent with previous non-model-based analysis presented in (McIntosh et al., 2024). **D,** Facilitation was strong in the targeted muscle and present in muscles innervated at nearby segments. ADM, abductor digiti minimi; APB, abductor pollicis brevis; ECR, extensor carpi radialis; FCR, flexor carpi radialis; PI, pairing interval.

## 4 Discussion

We tested the timing-dependent effects of paired brain and cervical SCS on arm and hand muscle responses in people living with chronic cervical SCI, those undergoing elective decompression for cervical myelopathy, and uninjured volunteers. We confirmed our hypothesis that synchronizing brain stimuli to converge with spinal cord stimuli at cervical levels facilitates upper extremity motor responses more strongly than stimuli arriving at non-convergent intervals. Facilitation occurred both in uninjured individuals and in people with spinal cord pathology, supporting the hypothesis that this pairing approach targets spared sensorimotor circuit interactions within the spinal cord both at and below the level of injury. No correlations were observed with severity or level of injury. Although the facilitatory effects of noninvasive paired stimulation are much lower in magnitude than those noted with epidural stimulation, we noted that increasing subthreshold spinal stimulation intensity produced greater effects. These observations will be useful in future applications of repetitive paired stimulation aimed to produce longer-lasting neural plasticity.

### 4.1 Significance of timing

Based on our prior evidence in animals and humans (McIntosh et al., 2024; Mishra et al., 2017; Pal et al., 2022, p. 20; Wecht et al., 2021), we calibrated the timing of brain and SCS to measure muscle responses to synchronous as compared to non-synchronous arrival of brain and spinal stimuli at the cervical spinal cord. The finding that facilitation was greatest at a PI of 0 (synchronous arrival) confirms the cervical spinal cord (as opposed to the brain or elsewhere) as the site of interaction in this pairing paradigm. More specifically, the dependence of facilitation on precise timing is consistent with the hypothesis that the interaction occurs between sensory afferents with descending motor connections in the cervical spinal cord.

### 4.2 Magnitude of effect in noninvasive versus invasive paired stimulation

Many factors may underlie the differences noted in the magnitude of facilitation between invasive and noninvasive paired stimulation:

#### 4.2.1. Precise localization

The ability to precisely localize the stimulation focal point in epidural as compared to noninvasive stimulation likely plays the largest of many factors contributing to the greater pairing effect with invasive pairing. Intraoperative stimulation during elective surgery in humans is delivered via two epidural electrode contacts with 1.3 mm cross-sectional area spaced 15 mm apart (McIntosh et al., 2024), as compared to the 50 cm cross-sectional area of the noninvasive cathode placed over the skin. Using precise epidural localization, stimulation slightly lateral to the midline produces much more facilitation than over the midline (Mishra et al., 2017). This degree of localization is not possible with TSCS. The broader field induced during noninvasive stimulation may trigger activity in multiple neural circuits, some of which may counteract and thereby reduce the resulting evoked potential. For example, non-specific excitation of sensory afferents across more than one cord segment may lead to coactivation of recurrent inhibitory circuits serving the target muscle and its antagonist(s). Although brain stimulation was noninvasive in both the operative and awake participants, the method used during surgery (transcranial electrical stimulation, TES) differs from TMS in at least two important ways: TES uses intradermal electrodes, thereby lowering skin impedance; and TES predominantly activates corticospinal motor neurons directly, as opposed to the indirect activation of corticospinal motor neurons by TMS (Terao & Ugawa, 2002). The latter difference results in more consistently timed descending cortical volleys, potentially enhancing precision of the pairing effect.

#### 4.2.2. Awake versus anesthetized state

Differences in the state of consciousness between participants in awake noninvasive versus anesthetized invasive experiments may greatly affect responses to brain stimulation. The most obvious effect would be increased variability in amplitude (and latency) of responses to brain stimulation in noninvasive experiments due to fluctuations in levels of alertness (Noreika et al., 2020; Ohtaki et al., 2017), as well as head movements in the awake state that may affect relative positioning between the TMS coil and scalp (Zorn et al., 2012). These sources of variability would decrease the signal-to-noise ratio of observed pairing effects.

#### 4.2.3. Stimulation timing

Prior invasively delivered paired stimulation experiments found peak facilitation at a consistent 9 ms ISI between brain and spinal stimulation (McIntosh et al., 2024), which corresponds to a PI of 0 (see Section 2.9). In our noninvasive human experiments, we have observed a high degree of variability in the latency of TMS responses and underlying CMCT, both within and across participants. Not surprisingly, CMCT was more consistent in uninjured individuals (ranging 6.3 to 9.9 ms) than in participants with SCI (ranging 6.1 to 23.4 ms). Rather than using a fixed *ISI* across participants, we therefore calibrated ISIs to focus on *PIs* centered around 0 (CMCT - 1.5 ms), indicating simultaneous convergence. The intra- and inter-participant variability of TMS response latency likely introduced subtle variations in CMCT-dependent PIs, thereby reducing the signal-to-noise ratio when measuring pairing effects.

### 4.3 Intensity effects

Our main hypothesis when testing the immediate effects of paired stimulation centered on the use of subthreshold TSCS to facilitate MEP response to suprathreshold TMS. Using suprathreshold TSCS would have two disadvantages: it would complicate distinguishing synergistic from additive effects, and it would be less tolerable for the intended use of repetitive spinal stimulation to achieve lasting effects.

We used varying methods to choose the spinal stimulation intensity with which to pair brain stimulation over the course of the project (section 2.7 and Table 1). In some cases, the stimulation intensity used during sessions was less than half of the hbMEP threshold. These variations in methodology, although a limitation in one sense, allowed us to discern differing pairing effects across a range of spinal stimulation intensities. Somewhat intuitively, we observed greater pairing effects when spinal stimulation intensity increased toward threshold (Fig. 4). This confirms the findings of our prior invasive and noninvasive studies (McIntosh et al., 2024; Wecht et al., 2021). Therefore, to maximize pairing effects in future research, we recommend applying spinal stimulation at intensities more consistently close to threshold.

### 4.4 Study Strengths and Limitations

Our main conclusion is that paired stimulation of motor cortex and spinal afferents results in facilitation that is strongly dependent on timing. We used analytical methods that allow comparison to our group’s past pairing experiments done invasively in people undergoing elective cervical spine surgery. We showed a significant facilitatory effect of pairing stimuli to arrive simultaneously within the cervical cord in comparison to non-convergent arrival. We also showed a clear relation between spinal stimulus intensity and facilitatory effect, which will help to optimize this pairing strategy.

The main weakness in our findings is the lower magnitude of facilitation noted using noninvasive paired stimulation in awake humans compared to invasive paired stimulation in anesthetized humans. The many potential reasons for this were discussed in section 4.2. Most importantly, it remains to be determined whether the statistically significant 10% overall facilitatory effect would translate into a clinically meaningful change when applied repetitively as a therapeutic modality. Our study was also limited by transitions in methodology for determining threshold and spinal stimulation intensity. However, this allowed us to better discern different facilitation effects with different applied spinal stimulation intensities. Furthermore, these experiments were not designed to identify the exact sites of interaction between brain and spinal cord stimulation. Future physiological studies in humans and animal models are necessary to elucidate the specific synaptic targets and mechanisms driving the observed pairing effect, as well as to understand the key factors underlying the differing magnitudes of facilitation seen between invasive and noninvasive approaches. Conclusively identifying these factors may provide modifiable targets for increasing the noninvasive pairing effect.

### 4.5 Implications for further research

Both non-invasive and epidural spinal stimulation facilitated cortical MEPs in a strongly timing-dependent manner. This suggests convergent mechanisms that can be used to compare efficacy for inducing plasticity and functional recovery. While the immediate effects of pairing are much stronger for epidural SCS compared with TSCS, the difference in lasting effects is not known. Other paired stimulation methods (Bunday et al., 2018; Bunday & Perez, 2012) rely on repetitive synchronized pairing to induce their effects, whereas immediate effects of single pairs have not been documented. Our previous experiments demonstrate that plasticity relies on proper timing of the two stimuli–no plasticity is induced with brain only, spinal only, or paired stimulation if it is inappropriately timed (e.g., 100 ms delay). Thus, the immediate effects experiments reported here will guide future plasticity experiments.

We intend to measure changes in paired stimulation facilitatory effect before, during, and after intervention in people who undergo elective cervical decompression surgery for spondylotic myelopathy. We further aim to apply paired stimulation repetitively with the goal of inducing lasting beneficial changes - an approach we term spinal cord associative plasticity (SCAP). We have demonstrated that SCAP applied repeatedly to rats with C4 contusion SCI increased spinal cord excitability and improved forelimb function (Pal et al., 2022) and we are currently working on similar approaches in humans using both operative and noninvasive techniques. Noninvasively, our vision involves combining repetitive paired facilitation with task-specific exercise, repurposed medications, or other approaches known to induce and consolidate beneficial plasticity in humans with spinal cord pathology.

## 5 Conclusion

This study provides compelling evidence that synchronizing brain and spinal cord stimulation to converge within the spinal cord facilitates upper extremity motor responses in both individuals with chronic cervical SCI and uninjured individuals. While the magnitude of this noninvasive facilitation effect is less than that observed with invasive epidural stimulation, the timing-dependent nature of the effect strongly suggests a conserved mechanism and highlights the cervical spinal cord as a key target for both invasive and noninvasive pairing. Future studies will determine ways to apply paired stimulation to induce long-lasting and clinically meaningful changes in individuals living with neurological conditions.

## Data Availability

The datasets generated during this study are available at: https://doi.org/10.5281/zenodo.15225065.

## 6 Acknowledgements

We express our sincerest thanks to all the participants for their time and involvement in the study. We extend this appreciation to Francisco E. Castano, Grace O. Famodimu, Kyla S. Holbrook, Lauren E. Kinne, Gregory A. Mendez, Joseph M. Robbins, William M. Savage, Caitlyn N. Sigafose, and Astrea Villarroel-Sanchez for their assistance in conducting experiments.

## 7 Credit author statement

Conceptualization: MSV, JBC, NYH; Data Curation: LMM, JRM, JAG, MXL; Formal Analysis: LMM, JRM, VT; Funding Acquisition: MSV, JBC, NYH; Investigation: LMM, JRM, JAG, MXL, SPS, EFJ, CM, MSV, VT, JBC, NYH; Methodology: LMM, JRM, JAG, VT, JBC, NYH; Project Administration: MSV, JBC, NYH; Resources: JBC, NYH; Software: JRM, VT, YKW; Supervision: LMM, JRM, JAG, MSV, JBC, NYH; Writing – Original Draft: LMM, JRM, JBC, NYH Writing – Review & Editing: LMM, JRM, JAG, EFJ, MSV, VT, JBC, NYH.

All authors have seen and approved the manuscript.

## 8 Funding

This study was funded by the National Institutes of Health (1R01NS124224); New York State Department of Health (C39070; N.Y.H. Investigator Initiated); and the Travis Roy Foundation Boston, MA (J.B.C. Investigator Initiated).

## 9 Competing interests

*Jason B. Carmel* is a Founder and stockholder in BackStop Neural and a scientific advisor and stockholder in SharperSense; he has received honoraria from Pacira, Motric Bio, and Restorative Therapeutics. *Michael S. Virk* has been a consultant and has received honoraria from Depuy Synthes, and stockholder with OnPoint Surgical. *Noam Y. Harel* is a consultant for RubiconMD. All other authors declare no potential conflicts of interest with respect to the research, authorship, and/or publication of this article.

## 10 Availability of data and materials

The datasets generated during this study are available at: https://doi.org/10.5281/zenodo.15225065.

## 12 Supplementary material

### 12.1 Recruitment curve fitting model

**Figure S1.**
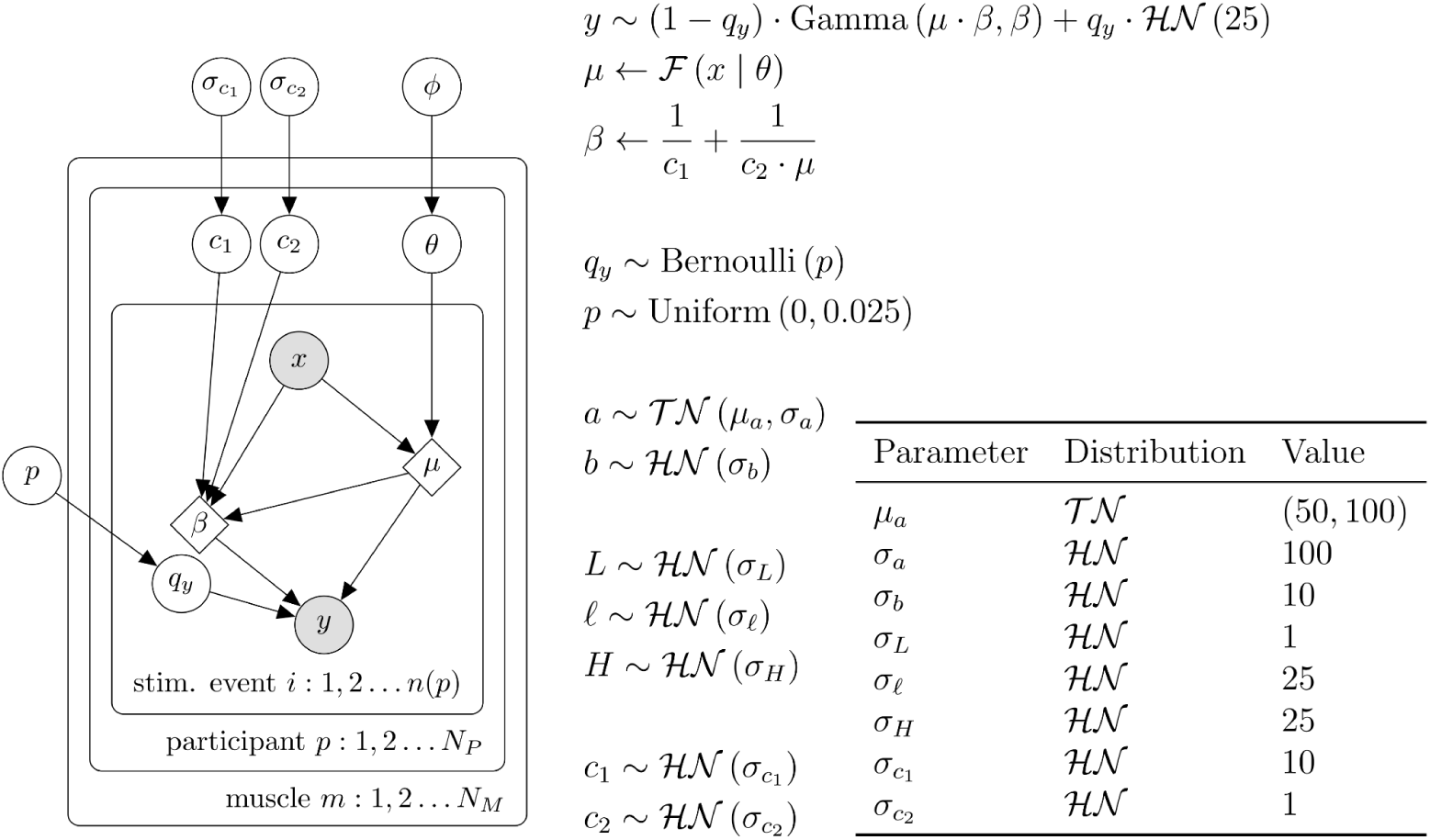
Recruitment curve model used for both TMS and TSCS. Here *F* is the rectified-logistic function, *θ*, and *θ* = {*a, b, L, ℓ, H*}, and *ϕ* = {*μ_a_, σ_a_, σ_b_, σ_L_, σ_ℓ_, σ_H_*}. The table gives the hyperpriors.

### 10.2 Facilitation is independent of injury level, severity, or chronicity

**Figure S2.**
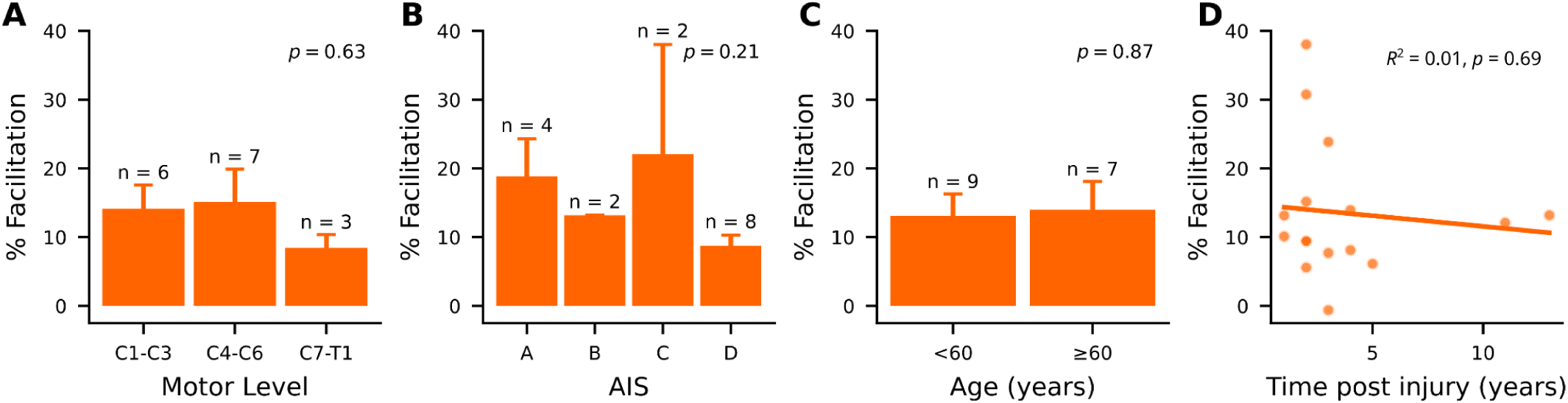
Facilitation is independent of injury level, severity, or chronicity. **(A–C)** Categorized variables relating to facilitation. Error bars represent SEM. **(A)** ANOVA indicated no significant effect of grouped motor level on facilitation (*p*-value shown in panel). **(B)** AIS classification did not significantly influence percent facilitation (ANOVA, *p*-value shown in panel). **(C)** Age was split into two similarly sized groups; ANOVA revealed no significant effect of age category on facilitation (ANOVA, *p*-value shown in panel). **(D)** Time post-injury was not significantly correlated with facilitation (linear regression, *p*-value shown in panel).

## Notes

### Clinical Trial

NCT05163639

### Funding Statement

This study was funded by the National Institutes of Health (1R01NS124224); the New York State Department of Health (N.Y.H. Investigator Initiated); and the Travis Roy Foundation Boston, MA (J.B.C. Investigator Initiated).

### Summary of Updates

Funding acknowledgments updated to include salary support from New York State Department of Health.

## References

1. Bunday, K. L., & Perez, M. A. (2012). Motor recovery after spinal cord injury enhanced by strengthening corticospinal synaptic transmission. Current Biology: CB, 22(24), 2355–2361. 10.1016/j.cub.2012.10.046

2. Bunday, K. L., Urbin, M. A., & Perez, M. A. (2018). Potentiating paired corticospinal-motoneuronal plasticity after spinal cord injury. Brain Stimulation, 11(5), 1083–1092. 10.1016/j.brs.2018.05.006

3. Bunge, R. P., Puckett, W. R., Becerra, J. L., Marcillo, A., & Quencer, R. M. (1993). Observations on the pathology of human spinal cord injury. A review and classification of 22 new cases with details from a case of chronic cord compression with extensive focal demyelination. Advances in Neurology, 59, 75–89.

4. Cortes, M., Thickbroom, G. W., Valls-Sole, J., Pascual-Leone, A., & Edwards, D. J. (2011). Spinal associative stimulation: A non-invasive stimulation paradigm to modulate spinal excitability. Clinical Neurophysiology: Official Journal of the International Federation of Clinical Neurophysiology, 122(11), 2254–2259. 10.1016/j.clinph.2011.02.038

5. Dixon, L., Ibrahim, M. M., Santora, D., & Knikou, M. (2016). Paired associative transspinal and transcortical stimulation produces plasticity in human cortical and spinal neuronal circuits. Journal of Neurophysiology, 116(2), 904–916. 10.1152/jn.00259.2016

6. Einhorn, J., Li, A., Hazan, R., & Knikou, M. (2013). Cervicothoracic multisegmental transpinal evoked potentials in humans. PloS One, 8(10), e76940. 10.1371/journal.pone.0076940

7. Fitzpatrick, S. C., Luu, B. L., Butler, J. E., & Taylor, J. L. (2016). More conditioning stimuli enhance synaptic plasticity in the human spinal cord. Clinical Neurophysiology: Official Journal of the International Federation of Clinical Neurophysiology, 127(1), 724–731. 10.1016/j.clinph.2015.03.013

8. Greiner, N., Barra, B., Schiavone, G., Lorach, H., James, N., Conti, S., Kaeser, M., Fallegger, F., Borgognon, S., Lacour, S., Bloch, J., Courtine, G., & Capogrosso, M. (2021). Recruitment of upper-limb motoneurons with epidural electrical stimulation of the cervical spinal cord. Nature Communications, 12(1), 435. 10.1038/s41467-020-20703-1

9. Guiho, T., Baker, S. N., & Jackson, A. (2021). Epidural and transcutaneous spinal cord stimulation facilitates descending inputs to upper-limb motoneurons in monkeys. Journal of Neural Engineering, 18(4), 046011. 10.1088/1741-2552/abe358

10. Harel, N. Y., & Carmel, J. B. (2016). Paired Stimulation to Promote Lasting Augmentation of Corticospinal Circuits. Neural Plasticity, 2016, 7043767. 10.1155/2016/7043767

11. Kakulas, A. (1988). The applied neurobiology of human spinal cord injury: A review. Paraplegia, 26(6), 371–379. 10.1038/sc.1988.57

12. Kakulas, B. A. (1999). A review of the neuropathology of human spinal cord injury with emphasis on special features. The Journal of Spinal Cord Medicine, 22(2), 119–124. 10.1080/10790268.1999.11719557

13. Knikou, M. (2017). Spinal Excitability Changes after Transspinal and Transcortical Paired Associative Stimulation in Humans. Neural Plasticity, 2017, 6751810. 10.1155/2017/6751810

14. Kruschke, J. (2014). Doing Bayesian Data Analysis: A Tutorial with R, JAGS, and Stan (2nd edition). Academic Press. 10.1016/B978-0-12-405888-0.00001-5

15. Leukel, C., Taube, W., Beck, S., & Schubert, M. (2012). Pathway-specific plasticity in the human spinal cord. The European Journal of Neuroscience, 35(10), 1622–1629. 10.1111/j.1460-9568.2012.08067.x

16. McIntosh, J. R., Joiner, E. F., Goldberg, J. L., Greenwald, P., Dionne, A. C., Murray, L. M., Thuet, E., Modik, O., Shelkov, E., Lombardi, J. M., Sardar, Z. M., Lehman, R. A., Chan, A. K., Riew, K. D., Harel, N. Y., Virk, M. S., Mandigo, C., & Carmel, J. B. (2024). Timing-dependent synergies between motor cortex and posterior spinal stimulation in humans. The Journal of Physiology, 602(12), 2961–2983. 10.1113/JP286183

17. McIntosh, J. R., Joiner, E. F., Goldberg, J. L., Murray, L. M., Yasin, B., Mendiratta, A., Karceski, S. C., Thuet, E., Modik, O., Shelkov, E., Lombardi, J. M., Sardar, Z. M., Lehman, R. A., Mandigo, C., Riew, K. D., Harel, N. Y., Virk, M. S., & Carmel, J. B. (2023). Intraoperative electrical stimulation of the human dorsal spinal cord reveals a map of arm and hand muscle responses. Journal of Neurophysiology, 129(1), 66–82. 10.1152/jn.00235.2022

18. Meunier, S., Russmann, H., Simonetta-Moreau, M., & Hallett, M. (2007). Changes in spinal excitability after PAS. Journal of Neurophysiology, 97(4), 3131–3135. 10.1152/jn.01086.2006

19. Mishra, A. M., Pal, A., Gupta, D., & Carmel, J. B. (2017). Paired motor cortex and cervical epidural electrical stimulation timed to converge in the spinal cord promotes lasting increases in motor responses. The Journal of Physiology, 595(22), 6953–6968. 10.1113/JP274663

20. Murray, L. M., & Knikou, M. (2017). Remodeling Brain Activity by Repetitive Cervicothoracic Transspinal Stimulation after Human Spinal Cord Injury. Frontiers in Neurology, 8, 50. 10.3389/fneur.2017.00050

21. Nielsen, J. F. (1996). Logarithmic Distribution of Amplitudes of Compound Muscle Action Potentials Evoked by Transcranial Magnetic Stimulation. Journal of Clinical Neurophysiology, 13(5), 423.

22. Noreika, V., Kamke, M. R., Canales-Johnson, A., Chennu, S., Bekinschtein, T. A., & Mattingley, J. B. (2020). Alertness fluctuations when performing a task modulate cortical evoked responses to transcranial magnetic stimulation. NeuroImage, 223, 117305. 10.1016/j.neuroimage.2020.117305

23. Ohtaki, S., Akiyama, Y., Kanno, A., Noshiro, S., Hayase, T., Yamakage, M., & Mikuni, N. (2017). The influence of depth of anesthesia on motor evoked potential response during awake craniotomy. Journal of Neurosurgery, 126(1), 260–265. 10.3171/2015.11.JNS151291

24. Pal, A., Park, H., Ramamurthy, A., Asan, A. S., Bethea, T., Johnkutty, M., & Carmel, J. B. (2022). Spinal cord associative plasticity improves forelimb sensorimotor function after cervical injury. Brain: A Journal of Neurology, 145(12), 4531–4544. 10.1093/brain/awac235

25. Pulverenti, T. S., Islam, M. A., Alsalman, O., Murray, L. M., Harel, N. Y., & Knikou, M. (2019). Transspinal stimulation decreases corticospinal excitability and alters the function of spinal locomotor networks. Journal of Neurophysiology, 122(6), 2331–2343. 10.1152/jn.00554.2019

26. Pulverenti, T. S., Zaaya, M., Grabowski, M., Grabowski, E., Islam, M. A., Li, J., Murray, L. M., & Knikou, M. (2021). Neurophysiological Changes After Paired Brain and Spinal Cord Stimulation Coupled With Locomotor Training in Human Spinal Cord Injury. Frontiers in Neurology, 12, 627975. 10.3389/fneur.2021.627975

27. Ridding, M. C., & Ziemann, U. (2010). Determinants of the induction of cortical plasticity by non-invasive brain stimulation in healthy subjects. The Journal of Physiology, 588(Pt 13), 2291–2304. 10.1113/jphysiol.2010.190314

28. Rossi, S., Antal, A., Bestmann, S., Bikson, M., Brewer, C., Brockmöller, J., Carpenter, L. L., Cincotta, M., Chen, R., Daskalakis, J. D., Di Lazzaro, V., Fox, M. D., George, M. S., Gilbert, D., Kimiskidis, V. K., Koch, G., Ilmoniemi, R. J., Lefaucheur, J. P., Leocani, L., … basis of this article began with a Consensus Statement from the IFCN Workshop on “Present, Future of TMS: Safety, Ethical Guidelines”, Siena, October 17-20, 2018, updating through April 2020. (2021). Safety and recommendations for TMS use in healthy subjects and patient populations, with updates on training, ethical and regulatory issues: Expert Guidelines. Clinical Neurophysiology: Official Journal of the International Federation of Clinical Neurophysiology, 132(1), 269–306. 10.1016/j.clinph.2020.10.003

29. Rossi, S., Hallett, M., Rossini, P. M., Pascual-Leone, A., & Safety of TMS Consensus Group. (2009). Safety, ethical considerations, and application guidelines for the use of transcranial magnetic stimulation in clinical practice and research. Clinical Neurophysiology: Official Journal of the International Federation of Clinical Neurophysiology, 120(12), 2008–2039. 10.1016/j.clinph.2009.08.016

30. Rossini, P. M., Barker, A. T., Berardelli, A., Caramia, M. D., Caruso, G., Cracco, R. Q., Dimitrijević, M. R., Hallett, M., Katayama, Y., & Lücking, C. H. (1994). Non-invasive electrical and magnetic stimulation of the brain, spinal cord and roots: Basic principles and procedures for routine clinical application. Report of an IFCN committee. Electroencephalography and Clinical Neurophysiology, 91(2), 79–92. 10.1016/0013-4694(94)90029-9

31. Sangari, S., Lundell, H., Kirshblum, S., & Perez, M. A. (2019). Residual descending motor pathways influence spasticity after spinal cord injury. Annals of Neurology, 86(1), 28–41. 10.1002/ana.25505

32. Schecklmann, M., Schmaußer, M., Klinger, F., Kreuzer, P. M., Krenkel, L., & Langguth, B. (2020). Resting motor threshold and magnetic field output of the figure-of-8 and the double-cone coil. Scientific Reports, 10(1), 1644. 10.1038/s41598-020-58034-2

33. Shulga, A., Zubareva, A., Lioumis, P., & Mäkelä, J. P. (2016). Paired Associative Stimulation with High-Frequency Peripheral Component Leads to Enhancement of Corticospinal Transmission at Wide Range of Interstimulus Intervals. Frontiers in Human Neuroscience, 10, 470. 10.3389/fnhum.2016.00470

34. Stefan, K., Kunesch, E., Benecke, R., Cohen, L. G., & Classen, J. (2002). Mechanisms of enhancement of human motor cortex excitability induced by interventional paired associative stimulation. The Journal of Physiology, 543(Pt 2), 699–708. 10.1113/jphysiol.2002.023317

35. Stefan, K., Kunesch, E., Cohen, L. G., Benecke, R., & Classen, J. (2000). Induction of plasticity in the human motor cortex by paired associative stimulation. Brain: A Journal of Neurology, 123 *Pt* *3*, 572–584. 10.1093/brain/123.3.572

36. Taylor, J. L., & Martin, P. G. (2009). Voluntary motor output is altered by spike-timing-dependent changes in the human corticospinal pathway. The Journal of Neuroscience: The Official Journal of the Society for Neuroscience, 29(37), 11708–11716. 10.1523/JNEUROSCI.2217-09.2009

37. Terao, Y., & Ugawa, Y. (2002). Basic mechanisms of TMS. Journal of Clinical Neurophysiology: Official Publication of the American Electroencephalographic Society, 19(4), 322–343. 10.1097/00004691-200208000-00006

38. Tyagi, V., Murray, L. M., Asan, A. S., Mandigo, C., Virk, M. S., Harel, N. Y., Carmel, J. B., & McIntosh, J. R. (2024). Hierarchical Bayesian estimation of motor-evoked potential recruitment curves yields accurate and robust estimates. *ArXiv*, arXiv:2407.08709v1.

39. Wecht, J. R., Savage, W. M., Famodimu, G. O., Mendez, G. A., Levine, J. M., Maher, M. T., Weir, J. P., Wecht, J. M., Carmel, J. B., Wu, Y.-K., & Harel, N. Y. (2021). Posteroanterior Cervical Transcutaneous Spinal Cord Stimulation: Interactions with Cortical and Peripheral Nerve Stimulation. Journal of Clinical Medicine, 10(22), 5304. 10.3390/jcm10225304

40. Wolters, A., Sandbrink, F., Schlottmann, A., Kunesch, E., Stefan, K., Cohen, L. G., Benecke, R., & Classen, J. (2003). A temporally asymmetric Hebbian rule governing plasticity in the human motor cortex. Journal of Neurophysiology, 89(5), 2339–2345. 10.1152/jn.00900.2002

41. Wu, Y.-K., Levine, J. M., Wecht, J. R., Maher, M. T., LiMonta, J. M., Saeed, S., Santiago, T. M., Bailey, E., Kastuar, S., Guber, K. S., Yung, L., Weir, J. P., Carmel, J. B., & Harel, N. Y. (2020). Posteroanterior cervical transcutaneous spinal stimulation targets ventral and dorsal nerve roots. Clinical Neurophysiology: Official Journal of the International Federation of Clinical Neurophysiology, 131(2), 451–460. 10.1016/j.clinph.2019.11.056

42. Yamashita, A., Murakami, T., Hattori, N., Miyai, I., & Ugawa, Y. (2021). Intensity dependency of peripheral nerve stimulation in spinal LTP induced by paired associative corticospinal-motoneuronal stimulation (PCMS). PloS One, 16(11), e0259931. 10.1371/journal.pone.0259931

43. Zorn, L., Renaud, P., Bayle, B., Goffin, L., Lebosse, C., de Mathelin, M., & Foucher, J. (2012). Design and Evaluation of a Robotic System for Transcranial Magnetic Stimulation. IEEE Transactions on Biomedical Engineering, 59(3), 805–815. IEEE Transactions on Biomedical Engineering. 10.1109/TBME.2011.2179938

